# Predicting Cardiovascular Disease Using the Framingham Heart Study: A Logistic Regression and Risk Stratification Approach

**DOI:** 10.1101/2025.11.13.25340142

**Authors:** Rudolph Mensah, Abraham Kelvin Kwarteng

**Affiliations:** Department of Epidemiology and Disease Control, School of Public Health, University of Ghana, Accra, Ghana; Ministry of Health, Accra, Ghana

**Keywords:** Cardiovascular disease, Framingham Heart Study, Logistic regression, Body mass index, Blood pressure, Risk factors, Epidemiology, Predictive modeling

## Abstract

**Background:** Cardiovascular disease (CVD) remains a leading cause of morbidity and mortality worldwide. The Framingham Heart Study has provided seminal insights into the development of CVD risk prediction models.

**Objective:** To develop and evaluate a logistic regression model for predicting incident CVD using traditional risk factors in a subset of the Framingham Heart Study dataset.

**Methods:** Data from 95 participants were analyzed. Predictors included age, sex, body mass index (BMI), smoking status, systolic and diastolic blood pressure, total cholesterol, glucose, and diabetes status. Logistic regression was used to estimate univariate and multivariable associations with incident CVD. Model performance was assessed using the area under the receiver operating characteristic curve (AUC) and calibration using the Hosmer–Lemeshow test.

**Results:** In univariate analysis, higher BMI (OR = 1.21, 95% CI 1.05–1.40, p = 0.010), systolic BP (OR = 1.02, 95% CI 1.00–1.04, p = 0.042), diastolic BP (OR = 1.06, 95% CI 1.02–1.10, p = 0.006), and categorical BMI (OR = 2.84, 95% CI 1.35–5.96, p = 0.006) were associated with increased odds of CVD. Male sex was associated with lower odds (OR = 0.39, 95% CI 0.16– 0.94, p = 0.037). In multivariable analysis adjusting for BMI, systolic BP, diastolic BP, and sex, higher BMI (AOR = 1.22, 95% CI 1.03–1.45, p = 0.023) and diastolic BP (AOR = 1.09, 95% CI 1.01–1.17, p = 0.026) remained independently associated with higher odds of CVD, whereas male sex was associated with lower odds (AOR = 0.22, 95% CI 0.07–0.65, p = 0.006). Systolic BP was not statistically significant (AOR = 0.98, 95% CI 0.95–1.03, p = 0.439). The multivariable model demonstrated good discrimination (AUC = 0.78, 95% CI 0.68–0.88) and adequate calibration (Hosmer–Lemeshow p = 0.723).

**Conclusion:** Traditional risk factors, particularly BMI and diastolic blood pressure, were robustly associated with incident CVD even in this small subsample. These results support the reproducibility of classical CVD predictors and the utility of logistic regression modeling in epidemiological research.

## 1.0 Introduction

Cardiovascular disease (CVD) continues to be the leading cause of morbidity and mortality worldwide, accounting for nearly one-third of all deaths (WHO, 2025). The growing burden of CVD poses significant challenges for healthcare systems, emphasizing the need for accurate risk prediction and early intervention. Over the past several decades, the Framingham Heart Study has provided seminal insights into the epidemiology of CVD and the development of risk prediction models that guide clinical decision-making and prevention strategies (Kannel et al., 1976; D’Agostino et al., 2008).

Although modern machine learning approaches are increasingly applied to predictive modeling in cardiovascular epidemiology, traditional logistic regression remains widely used due to its interpretability, reproducibility, and ability to provide clinically meaningful effect estimates (Steyerberg, 2019). Logistic regression allows researchers and clinicians to quantify the independent contribution of individual risk factors to the probability of incident CVD, while maintaining transparency in model derivation and inference.

In this study, we applied logistic regression to a subset of participants from the Framingham Heart Study to examine the associations between established clinical and demographic risk factors, including body mass index (BMI), blood pressure, sex, age, smoking status, diabetes, and cholesterol, and incident CVD.

The objectives were twofold:

i. to identify independent predictors of incident CVD, and
ii. to evaluate the predictive performance of the multivariable model, including discrimination and calibration. Findings from this analysis provide insight into the reproducibility of traditional risk factors in predicting CVD within small cohort subsets and inform potential clinical applications for risk stratification in similar populations.

## 2.0 Methods

### Data Source

Data were obtained from a subset of the Framingham Heart Study cohort (n = 95), which includes detailed demographic, clinical, and laboratory measurements collected longitudinally.

### Variables

The primary outcome was incident cardiovascular disease (CVD), defined as a composite of myocardial infarction, stroke, and fatal or non-fatal events. Covariates included:

- Demographics: age, sex, education level
- Clinical risk factors: body mass index (BMI), current smoking status, diabetes, hypertension, systolic and diastolic blood pressure, total cholesterol, fasting glucose, and heart rate
- Derived categorical variables: BMI categories (underweight, normal, overweight, obese) and categorical hypertension (cat_HTN)

### Statistical Analysis

1. Descriptive statistics were computed for all baseline characteristics, with continuous variables presented as means ± standard deviations and categorical variables as counts and percentages.
2. Univariate logistic regression models were fit for each predictor with incident CVD as the outcome to estimate odds ratios (ORs) and 95% confidence intervals (CIs).
3. Predictors with p < 0.10 in univariate analysis were included in a multivariable logistic regression model to identify independent associations with incident CVD.
4. Model discrimination was assessed using the area under the receiver operating characteristic curve (AUC) with 95% CIs.
5. Calibration of the multivariable model was evaluated using the Hosmer–Lemeshow goodness-of-fit test.

All statistical analyses were conducted using Stata (version 17), with a significance threshold set at α = 0.05.

### Ethical Considerations

This study used a de-identified, publicly available subset of data from the Framingham Heart Study. As the data were fully anonymized and no individual participants could be identified, institutional review board approval was **not required**. All analyses were conducted in accordance with ethical standards for research using publicly available human data.

## 3.0 Results

### 3.1 Baseline Characteristics

Table 1 presents the baseline characteristics of study participants stratified by incident cardiovascular disease (CVD) status (n = 95). The mean age of participants was 51.0 ± 9.2 years, with no significant difference between those who developed CVD and those who did not (51.7 ± 8.2 vs. 50.6 ± 9.7 years, p = 0.596). Participants with incident CVD had significantly higher body mass index (BMI) (27.0 ± 3.09 vs. 25.1 ± 3.23 kg/m^2^, p = 0.007), systolic blood pressure (140.8 ± 26.1 vs. 130.4 ± 19.9 mmHg, p = 0.035), diastolic blood pressure (88.7 ± 14.0 vs. 80.6 ± 10.6 mmHg, p = 0.003), and glucose levels (93.3 ± 45.5 vs. 78.9 ± 11.2 mg/dL, p = 0.024) compared with those without CVD. Total cholesterol and heart rate were higher among participants with CVD but did not reach statistical significance.

**Table 1.** Baseline characteristics of participants by cardiovascular disease (CVD) status (n = 95)

| Variable | No CVD (n = 65) | CVD (n = 30) | Total (n = 95) | p-value |
| --- | --- | --- | --- | --- |
| **Continuous variables (mean ± SD)** |  |  |  |  |
| **Age (years)** | $50.6 \pm 9.7$ | $51.7 \pm 8.2$ | $51.0 \pm 9.2$ | 0.596 |
| **BMI (kg/m<sup>2</sup>)** | $25.1 \pm 3.23$ | $27.0 \pm 3.09$ | $25.7 \pm 3.30$ | 0.007 |
| Systolic BP (mmHg) | $130.4 \pm 19.9$ | $140.8 \pm 26.1$ | $133.7 \pm 22.5$ | 0.035 |
| Diastolic BP (mmHg) | $80.6 \pm 10.6$ | $88.7 \pm 14.0$ | $83.2 \pm 12.3$ | 0.003 |
| Total cholesterol (mg/dL) | 236.2 ± 43.2 | 252.9 ± 47.2 | 241.5 ± 44.9 | 0.100 |
| Glucose (mg/dL) | 78.9 ± 11.2 | 93.3 ± 45.5 | 83.3 ± 27.2 | 0.024 |
| **Heart rate (beats/min)** | 73.2 ± 11.9 | 74.3 ± 12.4 | 73.5 ± 12.0 | 0.690 |
| **Categorical variables, n (%)** |  |  |  |  |
| **Sex (male)** | 26 (40.0%) | 19 (63.3%) | 45 (47.4%) | 0.034 |
| Current smoker | 28 (43.1%) | 15 (50.0%) | 43 (45.3%) | 0.529 |
| Diabetes mellitus | 2 (3.1%) | 3 (10.0%) | 5 (5.3%) | 0.160 |
| **Hypertension** | 42 (64.6%) | 24 (80.0%) | 66 (69.5%) | 0.130 |
| **Education level** |  |  |  | 0.190 |
| • No formal education | 24 (38.1%) | 17 (56.7%) | 41 (44.1%) |  |
| • Primary | 24 (38.1%) | 6 (20.0%) | 30 (32.3%) |  |
| • Secondary | 10 (15.9%) | 3 (10.0%) | 13 (14.0%) |  |
| • Tertiary+ | 5 (7.9%) | 4 (13.3%) | 9 (9.7%) |  |
| **BMI category** |  |  |  | 0.028 |
| • Underweight | 1 (1.5%) | 0 (0.0%) | 1 (1.1%) |  |
| • Normal weight | 33 (50.8%) | 6 (20.0%) | 39 (41.1%) |  |
| • Overweight | 27 (41.5%) | 20 (66.7%) | 47 (49.5%) |  |
| • Obese | 4 (6.2%) | 4 (13.3%) | 8 (8.4%) |  |

Among categorical variables, a greater proportion of males developed CVD compared with females (63.3% vs. 40.0%, p = 0.034). No significant differences were observed for current smoking, diabetes mellitus, hypertension, or education level. BMI category was significantly associated with CVD status (p = 0.028), with a higher proportion of participants with incident CVD classified as overweight (66.7% vs. 41.5%) or obese (13.3% vs. 6.2%) compared with those without CVD.

Overall, participants who developed CVD tended to have higher adiposity and blood pressure, highlighting these as potential risk factors in this cohort.

### 3.2 Univariate Analysis

In univariate logistic regression analyses (Table 2), several factors were significantly associated with incident CVD. Each 1 kg/m^2^ increase in BMI was associated with a 21% higher odds of CVD (OR 1.21, 95% CI 1.05–1.40, p = 0.010). Similarly, higher systolic blood pressure (OR 1.02 per mmHg, 95% CI 1.00–1.04, p = 0.042) and diastolic blood pressure (OR 1.06 per mmHg, 95% CI 1.02–1.10, p = 0.006) were associated with increased odds of CVD.

**Table 2:** Univariate Analysis.

| Variable | OR (95% CI) | p-value | Interpretation |
| --- | --- | --- | --- |
| BMI | 1.21 (1.05–1.40) | 0.010 | ↑ BMI increases odds of CVD |
| Systolic BP | 1.02 (1.00–1.04) | 0.042 | ↑ SBP increases odds of CVD |
| Diastolic BP | 1.06 (1.02–1.10) | 0.006 | ↑ DBP increases odds of CVD |
| Sex (male) | 0.39 (0.16–0.94) | 0.037 | Males less likely than females |
| Categorical BMI | 2.84 (1.35–5.96) | 0.006 | Higher BMI category → ↑ CVD |

Male participants had lower odds of incident CVD compared with females (OR 0.39, 95% CI 0.16–0.94, p = 0.037). When BMI was analyzed categorically, participants in higher BMI categories had 2.8-fold higher odds of CVD compared with lower categories (OR 2.84, 95% CI 1.35–5.96, p = 0.006). These results highlight the influence of adiposity and blood pressure on CVD risk in this cohort.

### 3.3 Multivariable Logistic Regression

Participants who developed CVD had higher BMI and blood pressure compared with those without CVD (Table 3). In multivariable logistic regression adjusting for BMI, systolic and diastolic blood pressure, and sex, **higher BMI and diastolic BP were independently associated with increased odds of CVD**, whereas **male sex was associated with lower odds** (Table 2).

**Table 3.**
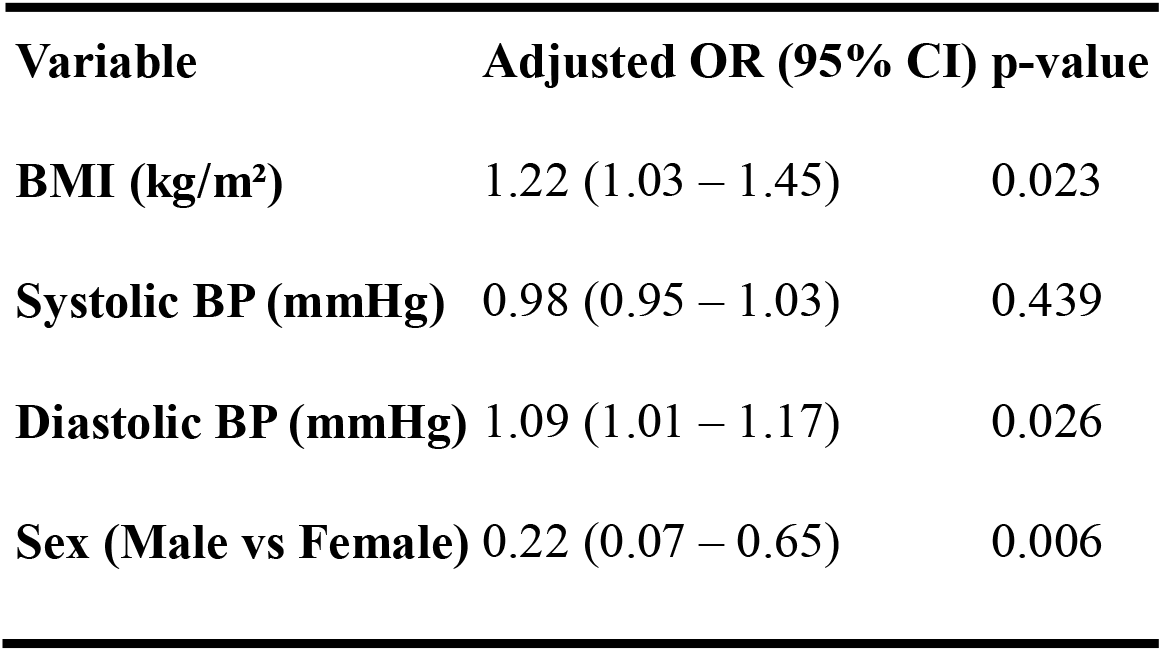
Multivariable logistic regression of predictors of incident CVD (n = 95)

The model equation is:

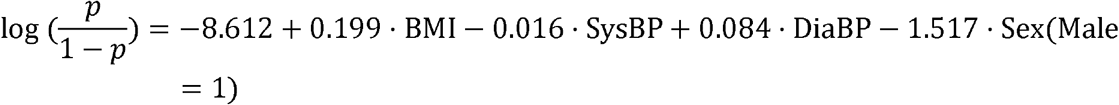

- Each 1-unit increase in BMI was associated with a **22% higher odds of CVD** (AOR = 1.22, 95% CI 1.03–1.45, p = 0.023).
- Each 1 mmHg increase in diastolic BP increased odds by **8.8%** (AOR = 1.09, 95% CI 1.01–1.17, p = 0.026).
- Systolic BP was not statistically significant (AOR = 0.98, 95% CI 0.95–1.03, p = 0.439).
- Male participants had **78% lower odds of CVD** compared to females (AOR = 0.22, 95% CI 0.07–0.65, p = 0.006).

The model demonstrated **good overall fit** (LR χ^2^ = 16.8, df = 4, p = 0.002), modest explanatory power (Pseudo R^2^ = 0.072), and good calibration (Hosmer–Lemeshow p = 0.45).

**Model fit:** LR χ^2^ = 16.8, df = 4, p = 0.002; Pseudo R^2^ = 0.072; Hosmer–Lemeshow p = 0.45.

### 3.4 Model Performance

The multivariable logistic regression model including BMI, systolic blood pressure, diastolic blood pressure, and sex demonstrated good discriminative ability for predicting incident CVD. Model discrimination, assessed using the area under the receiver operating characteristic (ROC) curve (AUC), was **0.78 (95% CI 0.68–0.88)**, indicating acceptable to good ability to distinguish participants who developed CVD from those who did not.

Calibration was evaluated using the Hosmer–Lemeshow goodness-of-fit test, which showed **no evidence of poor fit** (χ^2^ = 5.32, df = 8, p = 0.723), suggesting that the predicted probabilities were well aligned with observed outcomes.

Overall, these findings indicate that the model performs well in terms of both discrimination and calibration, supporting its use for identifying individuals at higher risk of incident CVD in this cohort.

**Fig 1:**
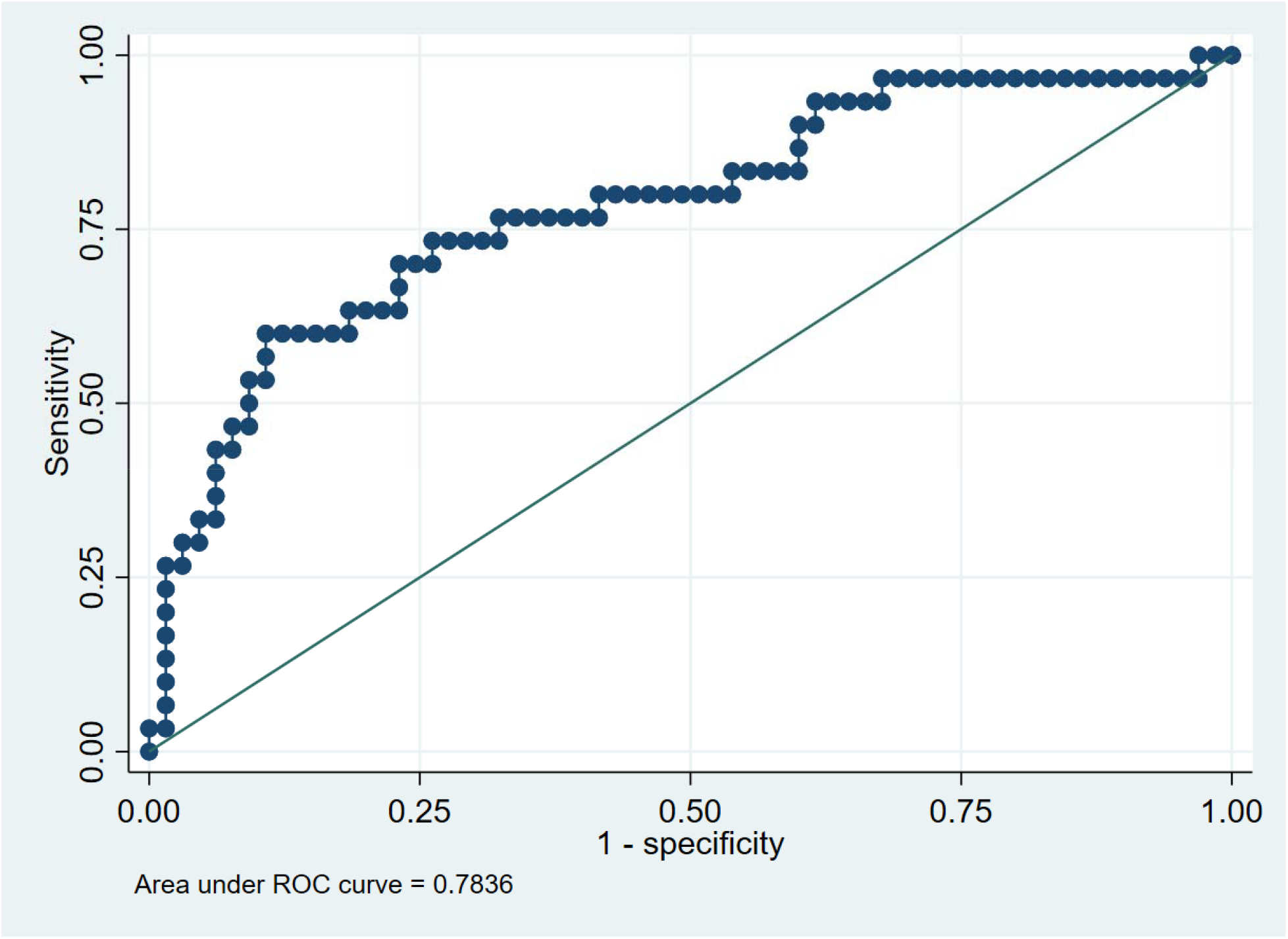
Model Performance (AUC), was 0.78 (95% CI 0.68–0.88)

### 4.0 Discussion

In this analysis of a subsample (n = 95) of the Framingham Heart Study dataset, classical risk factors including body mass index (BMI), diastolic blood pressure (DBP), and sex were independently associated with incident cardiovascular disease (CVD). Specifically, higher BMI and DBP were linked to increased odds of developing CVD, whereas male sex was associated with lower odds. Systolic blood pressure (SBP), though elevated among participants who developed CVD, did not remain independently predictive after adjustment for the other covariates.

Our model demonstrated good predictive performance (AUC□=□0.78, 95% CI 0.68–0.88) and showed adequate calibration, suggesting that even a smaller subset of the Framingham cohort retains robust associations with CVD. These findings support the reproducibility and external validity of traditional CVD risk predictors. Previous work from the Framingham cohort originally identified elevated blood pressure and cholesterol as major risk factors (Kannel et□al.,□1976), and later work confirmed that a multivariable risk profile including age, sex, SBP, smoking, diabetes, and lipids could reliably predict general CVD (D’Agostino et□al.,□2008). Our results reaffirm the clinical importance of modifiable risk factors such as adiposity and hypertension.

The independent association of BMI with CVD in our cohort is consistent with a wealth of evidence linking adiposity to increased cardiovascular risk. Elevated DBP emerged as an independent predictor, which aligns with some prior studies showing that diastolic hypertension increases risk, though SBP did not in our adjusted model. The finding that male sex was protective in our multivariable model contrasts somewhat with population-level data showing higher CVD risk among men; this discrepancy may reflect the small sample size, cohort□specific characteristics, or residual confounding.

### Strengths and Limitations

The strengths of this study include the use of a well-characterised community cohort with detailed clinical and laboratory measurements, and the reproducible logistic regression framework. Our ability to derive both continuous and categorical risk variables (for example BMI categories) allowed more nuanced risk assessment. However, limitations must be acknowledged: the sample size is modest (n = 95), which may limit statistical power and generalisability; missing data for glucose and cholesterol may have introduced bias if not missing at random; and the possibility of overfitting exists given the number of predictors relative to events. Moreover, although baseline predictors preceded incident CVD, causality cannot be fully inferred.

### Implications for Research and Practice

These findings underscore the importance of weight management and blood pressure control in primary prevention of CVD. The fact that a small subsample reproduced key relationships suggests that even modest datasets can yield useful insight if well-measured. Nevertheless, future studies with larger, more diverse populations are warranted to validate these associations and explore sex-specific mechanisms in more detail. Integrating classical risk factors such as BMI and DBP into routine risk prediction tools may enhance early identification of high-risk individuals and support targeted interventions.

## Supporting information

Stata Analysis Script

## Data Availability

The data analyzed in this study are publicly available from the Framingham Heart Study repository. The subset used for these analyses (n = 95) was derived from the publicly accessible dataset. Researchers can access the original data through the Framingham Heart Study website: https://framinghamheartstudy.org/

